# Association Between Entomological Indices and Malaria Test Positivity Rates in Western Kenya: Implications for Surveillance

**DOI:** 10.64898/2026.02.03.26345457

**Authors:** Brenda Omala, David Mburu, Maurice Ombok, Vincent Moshi, John Gimnig, Nicole L. Achee, John P. Grieco, Bernard Abong’o, Eric Ochomo

**Affiliations:** Pwani University Bioscience Research Centre (PUBReC), Pwani University, Kilifi, Kenya; Center for Global Health Research, Kenya Medical Research Institute, Kisumu, Kenya; Division of Parasitic Diseases and Malaria, Centre for Global Health, Centre for Disease Control and Prevention, Atlanta, GA, 30333, USA; Department of Biological Sciences, Eck Institute for Global Health, University of Notre Dame, Notre Dame, USA; Vector Group, Liverpool School of Tropical Medicine, Liverpool, UK; Centre for Infectious and Parasitic Disease Control Research, Kenya Medical Research Institute, Busia, Kenya

## Abstract

The relationship between malaria entomological indices and epidemiological outcomes remains poorly understood at local scales in western Kenya, limiting evidence-based surveillance strategies. This study evaluated associations between vector densities, entomological inoculation rates (EIR), human biting rates (HBR), and malaria test positivity rates in a high-transmission setting. The study was conducted in five villages within Teso South sub-County, Busia County, Kenya, from March to June 2021. Village-level entomological indices including vector densities, human biting rates (HBR), and entomological inoculation rates (EIR) were calculated for *Anopheles funestus* and *An. gambiae s.l*. Concurrent malaria test positivity rates were obtained from outpatient department registers in the health facilities serving the five villages (n=2,377 cases). The relationship between entomological indices and malaria test positivity was analyzed using multilevel logistic regression models, accounting for individual- and village-level factors. During this period 1,957 female anopheline mosquitoes, predominantly *An. gambiae s.l* (89.17%) and *An. funestus* (10.48%) were collected. Malaria test positivity rates varied significantly across villages (range: 23.6%-68.7%). Multilevel analysis revealed significant associations between malaria test positivity and both anopheline mean density (OR 1.12, 95% CI 1.08-1.16, P<0.0001) and HBR (OR 1.37, 95% CI 1.19-1.57, P<0.0001). Age of humans showed a slight negative association with malaria positivity (OR 0.999, 95% CI 0.998-0.9997, P=0.013), while EIR showed no significant association (OR 1.04, 95% CI 0.96-1.13, P=0.319). Entomological indices, particularly vector density and human biting rate, showed strong associations with malaria test positivity rates. However, entomological monitoring is actually pretty intensive and covers only a few villages, and only parts of those, while routine surveillance for clinical cases of malaria is becoming increasingly reliable. These findings therefore suggest that one solution to higher malaria cases is target transmission through vector control. This information could be valuable for national malaria control programs in optimizing surveillance strategies and evaluating intervention effectiveness.

## BACKGROUND

Despite significant advances in control measures, malaria remains a critical global public health challenge, particularly in sub-Saharan Africa. In the most recent World Malaria Report, there were an estimated 263 million malaria cases in 2023, with approximately 597,000 deaths globally [1]. In Kenya, while national prevalence was 5.6% according to the 2020 Kenya Malaria Indicator Survey, significant regional variations persist, with Busia County reporting a prevalence of 38%[2]. Effective malaria control requires robust surveillance systems that can accurately monitor both entomological and epidemiological indicators [3]. However, these two aspects of surveillance are often conducted independently, creating potential gaps in our understanding of transmission dynamics. While epidemiological surveillance through health facility data provides valuable information about disease burden, it often suffers from reporting inconsistencies and coverage limitations. Entomological surveillance, though more resource-intensive, can provide early warnings of transmission risk and intervention effectiveness and can identify failure of vector control interventions due to the quality of implementation, insecticide resistance, and behavioral changes in the mosquito vector [4].

The World Health Organization’s Global Vector Control Response (2017-2030) emphasizes surveillance as a crucial pillar for malaria elimination efforts [5]. National malaria control programs (NMCPs) conduct parallel but separate tracking of entomological and epidemiological indicators. These form the basis for evaluation of control interventions, monitoring temporal and spatial heterogeneity of *Plasmodium* parasitemia as well as malaria vectors [6]. Epidemiological data is often routinely collected passively through health facility records complemented with Malaria Indicator Surveys conducted once every 5 years. Entomology surveillance is conducted on a less regular, much smaller scale and often purposively for research or surveillance purposes, which is rarely on a large enough spatial scale to make an informed decision about wide administrative areas [7] unless powered sufficiently to do so [8]. This separation raises important questions about the relationship between these indicators and whether one set could effectively predict the other, potentially optimizing resource allocation in surveillance programs.

Since the entomological and epidemiological datasets are usually collected by independent groups at the NMCPs, efforts to understand their relatedness are rare. For new vector control paradigms, WHO requires evidence of epidemiological efficacy in at least two countries [9], with less emphasis on entomological outcomes, yet entomological data can be scaled up quite easily with the use of community health promoters [10]. Additionally, entomological data often include surveys of interventions’ coverage and use and can inform inherent gaps in coverage in targeted areas or regions. Moreover, recent evidence suggests that entomological data collection can be decentralized, and community-based teams trained to deploy traps and subsequent data at minimal cost [11].

While previous studies have examined relationships between individual indices and transmission [12] [13], few have comprehensively analyzed multiple entomological indicators simultaneously with epidemiological outcomes. Three key knowledge gaps remain: (1) the predictive value of multiple entomological indices in combination, (2) the spatial heterogeneity of these relationships at village level, and (3) the cost-effectiveness of different surveillance strategies in resource-limited settings. This study investigated the relationship between key malaria entomological indices (vector densities, entomological inoculation rates, and human biting rates) and malaria test positivity rates in five villages within Busia County, western Kenya. We hypothesized that entomological indices would show significant associations with malaria test positivity rates, potentially serving as predictive indicators for transmission intensity. Our findings could inform more efficient and integrated approaches to malaria surveillance and response, particularly in resource-limited settings.

## MATERIALS AND METHODS

### Study Design and Study Population

This study was nested within a cluster randomized control trial (cRCT) evaluating the epidemiological impact of spatial repellent’s impact on malaria incidence in western Kenya [14]. The study was carried out between March and June 2021, corresponding to the long rainy season, in five villages within Teso South sub-County, Busia County: Adungosi, Aciit, Kwangamor, Koruruma, and Okaereret (Fig 1). These villages were selected based on completeness of health facility data. Epidemiological data were obtained from the health facility records in the specific villages where mosquito collection was performed for correlation purposes. The study was conducted during the baseline period prior to the deployment of spatial repellents.

**Figure 1:**
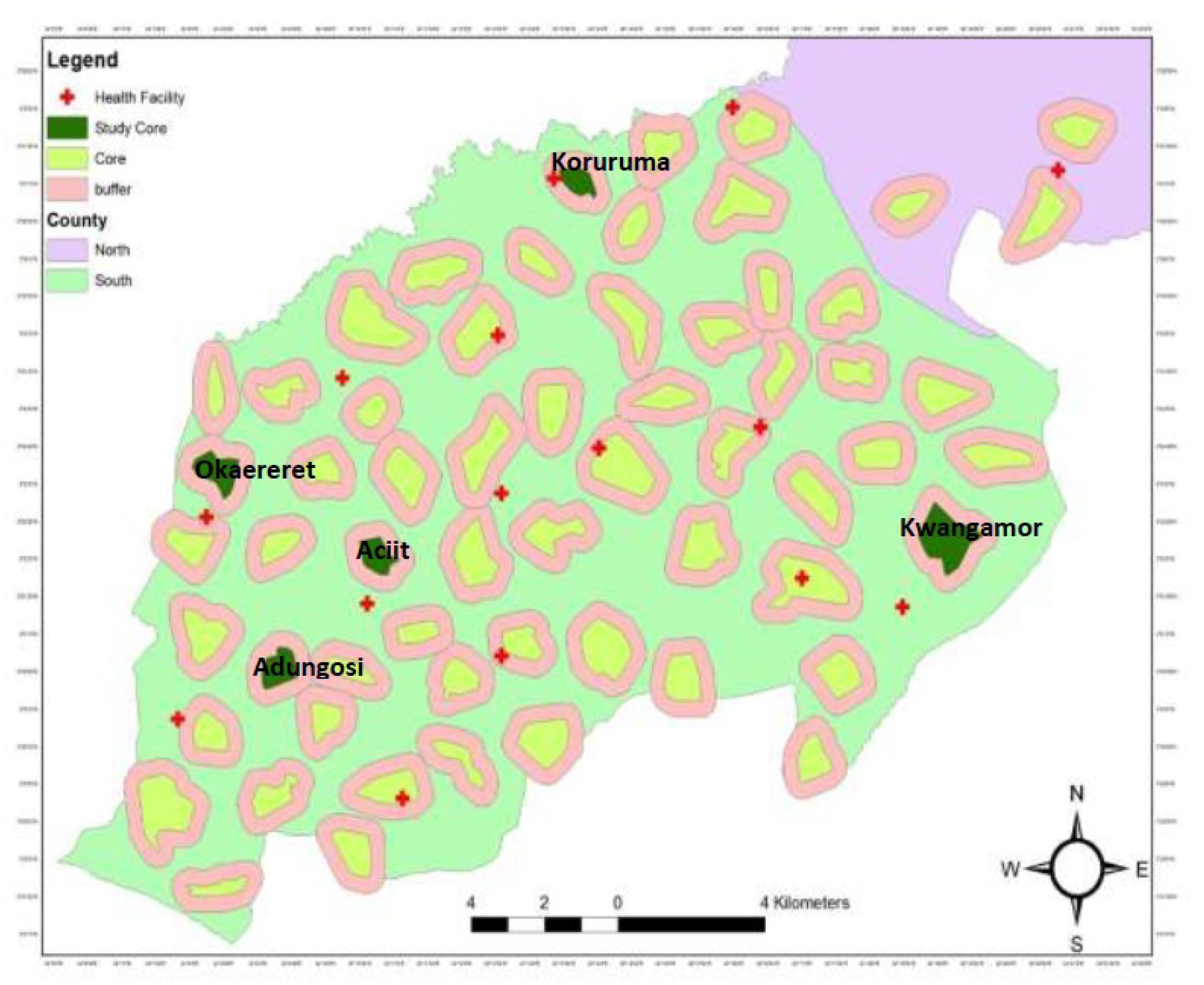
A map showing study sites in Teso South sub counties.

The study area is characterized by moderate to high rainfall (760-2000 mm annually) distributed across two seasons: long rains (March-May) and short rains (October-November). Busia County, situated within the Lake Victoria region, reports the highest malaria prevalence (38.5%) in Kenya. The region’s primary economic activities include subsistence farming and cash crop cultivation (sugarcane and cassava). Insecticide-treated nets (ITNs) are the primary vector control intervention in the area.

### Eligibility criteria

We included all cases that were from the five study villages and with complete variables of interest including patient’s identity number, test results, gender, reporting month and village of residence. Cases that failed to meet any of the mentioned parameters were excluded in the analysis.

### Sample size and sampling strategy

Sample size determination followed the protocol of the parent spatial repellent trial. For entomological surveillance, we randomly selected 10 households per village per month (total n=198 sampling events) using a computerized random number generator. For epidemiological data, we analyzed all available malaria test records from the five health facilities serving these villages during the study period (n=2,377 records).

### Health Facility Surveillance

Outpatient department data (OPD) from five health facilities adjacent (within 5km) to the study villages was retrieved and used to extract records of individuals coming from the study villages. Five facilities; Akiriamas Dispensary, Amaase Dispensary, Kwangamor Dispensary and Lukolis Model Health Center, participated in this study. Data was abstracted from ministry of health (MoH) OPD under five (204 A) and over five (204 B) registers of the chosen hospitals for similar months as of entomological data collection. A smart phone camera was used to photograph every page of the registers, and the images were converted into portable document formats (PDFs. Patients’ confidentiality was ensured by covering the column containing their names and other personal identifiers when taking photographs. The information was then recorded in Microsoft Excel.

### Outcome Measures from the Health Registers

The variables of interest for the study included: name of health facility (HF), age, gender, month, malaria test results. Malaria test positivity rate was determined as the fraction of persons diagnosed with malaria, over the overall number of individuals presenting at the health facility for each study village. The tests were conducted by microscopy technique or rapid diagnostic tests (mRDT). To facilitate linkage of the HF’s data to the respective village, patients’ area of residence was also abstracted as recorded on the OPD registers, and the outcome measures were calculated per village.

### Vector Surveillance

Mosquito collection was conducted using CDC light traps indoors in five villages between March and June 2021. Data from the indoor placed traps were used to estimate human biting rates (HBR) [15]. Ten (10) households were randomly selected in every village each month and used as study units for mosquito sample collection. Other metadata collected alongside the entomology questionnaire included: whether eaves were open or closed, type of roof and wall, information on cooking in the rooms as well as utilization of other mosquito control methods for instance mosquito coils, the night before collection. The traps were installed at 1800 hours in the evening, left to run all night and collected at 0700 the following morning. Trapped mosquitoes were then moved to site laboratory in trap cups for morphological characterization and further analyses.

### Identification of Anopheles mosquito species and sporozoites detection

Mosquito samples collected from the selected villages were classified by applying taxonomical keys of Coetzee [16]. Legs, wings and abdomen were cut from the mosquitoes to be utilized in polymerase chain reaction (PCR) examination [17] purposed for species level identification of members of *An. gambiae* Complex and *An. funestus s.l* group. Molecular identification was done on only up to a hundred samples per village per month. Sporozoite infection rate was conducted on heads and thoraces of the same selection of mosquitoes per village used for PCR species identification. Sporozoites were detected using sandwich Enzyme linked immunosorbent assay (ELISA) as described by prior studies [18].

### Data Analysis

The analytical approach focused on examining the relationship between entomological indices and malaria test positivity rate while accounting for both individual and village-level variations. All analyses were conducted using R statistical software (version 4.2.2), employing a systematic multilevel modelling approach to handle the nested structure of the data. To prepare the data for analysis, the distribution properties were examined. The Shapiro-Wilk test revealed ununiform distribution patterns in the key variables (P < 0.05), leading to the employment of non-parametric methods where appropriate. The Kruskal-Wallis test was used to examine differences in mosquito means between villages, as this test does not assume normal distribution.

### Determining Malaria Vector Mean Densities

Vector abundance was assessed using descriptive statistics (means, proportions, and 95% CI). The quotient from malaria vectors collected and number of households where the mosquitoes were collected in a village was considered as malaria vector density. This indicator gives a general view of information on malaria vector density per house in a village.

### Determining Sporozoites Infectivity (SR)

Sporozoite rates were derived by the formula given below: -

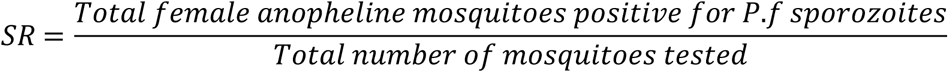

### Determining Human Biting Rates

Direct calculation approach was made from female anopheles mosquitoes collected using indoor CDC LTs and the number of sleepers inside a mosquito net with an assumption that the mosquitoes were seeking the human host under the bed net next to which the light trap was hung (WHO, 2013). The analysis was done per village using formula as shown below:

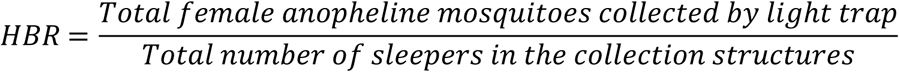

### Determining Entomological Inoculation Rate (EIR)

Entomological inoculation rate was obtained by multiplying sporozoites rates, human biting rates using a time period (4 months/122 days) as guided by WHO and other studies [19], [20].The EIR was calculated as follows: -

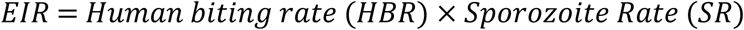

### Association Between Entomological and Epidemiological Indicators

The main analysis utilized multilevel logistic regression models to account for the hierarchical nature of the data, where individuals (level 1) were nested within villages (level 2). Individual ages were centred around the grand mean to facilitate interpretation and reduce multicollinearity. For village-level measures, the researchers created non-self-aggregate values, excluding each individual’s data point from their respective village-level calculations. The modelling process proceeded through four sequential stages: In the first stage, an empty model (null model) containing only the random intercept for villages was fitted. This calculation of the intraclass correlation coefficient (ICC) revealed that 14% of the variance in malaria test positivity was attributable to village-level differences, providing justification for the multilevel modelling approach. The second stage involved construction of a constrained intermediate model incorporating all individual-level variables (age, gender) and village-level variables (vector densities, HBR, EIR). This model elucidated the main effects of predictors while accounting for the nested data structure. In the third stage, the research team developed an augmented intermediate model incorporating random slopes for individual-level variables. Comparison with the constrained model using likelihood ratio tests determined whether allowing individual-level variable effects to vary across villages improved model fit. The final stage involved fitting a conclusive model with cross-level interactions between age and the entomological predictors (mean densities, HBR, and EIR). This analysis tested whether entomological indices’ effects on malaria test positivity varied with individual age. For all models, odds ratios with 95% confidence intervals we calculated, considering P-values less than 0.05 as statistically significant. Model fit was assessed using deviance statistics, and residuals were examined for model assumption violations.

Sensitivity analyses were conducted through: 1) the removal of influential observations and reassessment of key relationships, 2) testing of alternative correlation structures and 3) examination of missing data impact on conclusions. All statistical tests employed a significance level of α = 0.05. Model assumptions were verified through residual plots and influence diagnostics. Result reporting included measures of uncertainty (confidence intervals) alongside point estimates to provide comprehensive understanding of finding precision.

This approach allowed for addressing the hierarchical data structure, control for potential confounding at both individual and village levels, examination of cross-level interactions, generation of robust estimates of relationships between entomological indices and malaria test positivity and quantification of estimate uncertainty. The analysis provided the foundation for understanding the relationship between entomological indices and malaria transmission patterns in the study area, while accounting for the complex nature of the data structure.

### Ethical Consideration

Ethical clearance to conduct the study was obtained from the Kenya Medical Research Institute – Scientific and Ethics Review Unit (KEMRI-SERU) number 3870 and Pwani University Institutional Scientific and Ethics Review Committee, approval number ISERC/MSc/048/2022. During routine mosquito collection by CDC-LT verbal consents were obtained from household heads. All methods were performed according to stipulated guidelines and regulations.

## RESULTS

### Malaria Test Positivity Rates

The study findings were derived from a comprehensive analysis of both entomological and epidemiological data collected from five villages in Busia County, western Kenya. A total of 18,299 facility records were abstracted from the Ministry of Health (MOH) under five and over five years facility outpatient department registers. Of these 2,377 malaria test records met the inclusion criteria for analysis. Of those who were tested for malaria, 61.8% (n=1,470) were females and 38.2% (n=907) were males. The age distribution was: <5 years: 25.3% (n=601), 5-14 years: 35.7% (n=849), 15-45 years: 28.4% (n=675) and >45 years: 10.6% (n=252). Mean age was 16.8 years (SD: 15.4). The distribution of cases varied across the study villages, with the highest number of records being documented in Koruruma (n=915), followed by Kwangamor (n=690), while fewer cases were recorded in Okaereret (n=211), Aciit (n=311), and Adungosi (n=250).

More females (60%) than males (40%) were positive for malaria. School going children aged between 5-14 years had the highest malaria test positivity rate while children under five years and individuals above 14 years in age had the lowest test positivity rates. An overall malaria test positivity rate of 54.3% was calculated across all study sites. Significant heterogeneity in positivity rates was observed among villages, with the highest rate being recorded in Kwangamor (68.7%), followed by Okaereret (64.5%) and Koruruma (56.1%). Substantially lower rates were documented in Aciit (35.0%) and Adungosi (23.6%) (Table 1).

**Table 1:** Distribution of records and malaria cases in the study villages.

| Village | Total Records | Malaria Positive Cases | Malaria Test Positivity Rate (%) |
| --- | --- | --- | --- |
| Koruruma | 915 | 513 | 56.1 |
| Kwangamor | 690 | 474 | 68.7 |
| Okaereret | 211 | 136 | 64.5 |
| Aciit | 311 | 109 | 35.0 |
| Adungosi | 250 | 59 | 23.6 |
| **Total** | **2377** | **1291** | **54.3** |

Pairwise comparisons of the malaria test positivity rates between the study villages were conducted using Marascuilo Procedure (Table 2). The test showed significant difference between the village proportions apart from Aciit and Adungosi, Koruruma and Okaereret, Kwangamor and Okaereret.

**Table 2:**
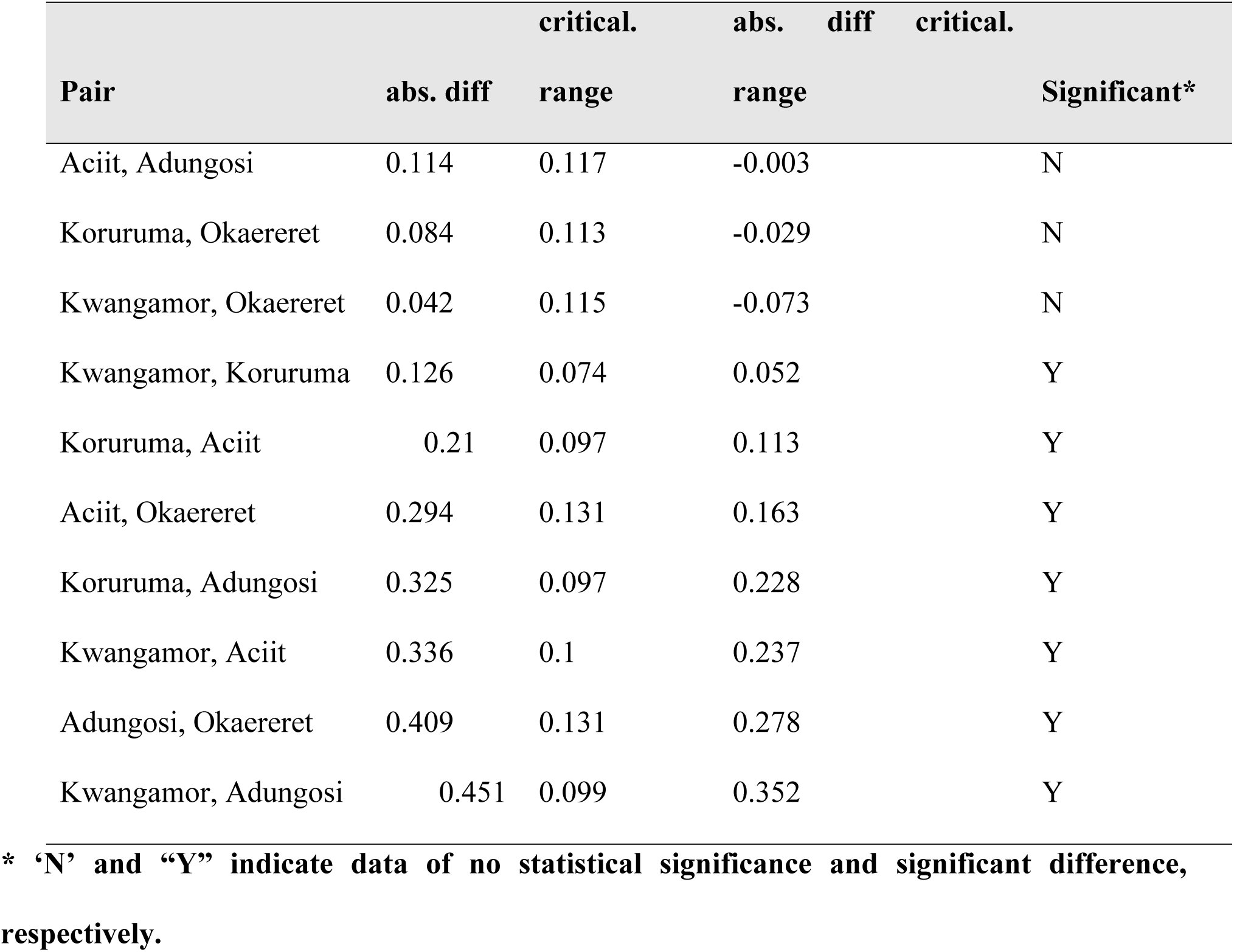
Marascuilo procedure for pair-wise comparison of malaria test positivity rates between study villages.

### Total Mosquitoes Collected

In the entomological surveillance, 1,957 female anopheline mosquitoes were collected and identified. The predominant species was *An. gambiae s.l*., comprising 89.2% (n=1,745) of the total collection, followed by *An. funestus* at 10.5% (n=205) (Table 4). A proportion of the *Anopheles* mosquitoes (n=661) was also identified by PCR and *An.gambiae s.s* was the predominant member of the An. gambiae complex (78.2%) (Table 3). The overall vector density across the study area was calculated at 10.0 mosquitoes per house per night, although marked variations were observed between villages. The highest densities were recorded in Kwangamor, Koruruma, and Okaereret, where 14.7, 13.6, and 11.6 mosquitoes per house per night were observed respectively. Lower densities were observed in Aciit and Adungosi, where only 2.8 and 1.5 mosquitoes per house per night were recorded (Table 3).

**Table 3:**
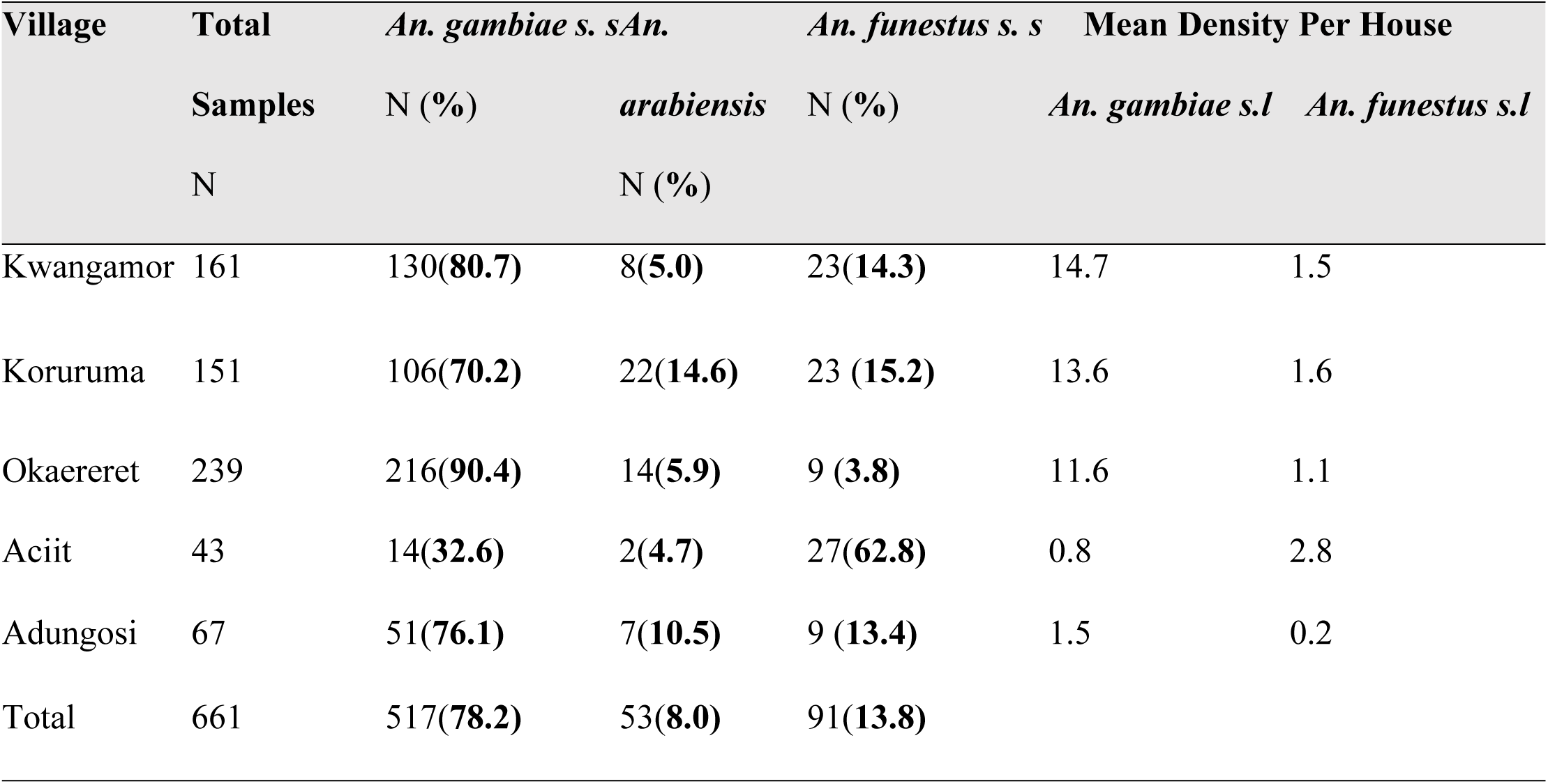
Proportion of *Anopheline* species by PCR and Anopheline species mean distribution per village.

**Table 4:** HBR for the study villages of Kwangamor, Koruruma, Okaereret, Aciit, Adungosi.

| Village | Total<br>collection | *An.gambiae s.l* | *An.funestus s.l* | HBR | EIR |
| --- | --- | --- | --- | --- | --- |
| Okaereret | 508 | 464(91%) | 44(9%) | 5.9 | 19.1 |
| Koruruma | 606 | 542(89%) | 64(11%) | 5.2 | 6.6 |
| Kwangamor | 630 | 572(91%) | 58(9%) | 4.2 | 1.5 |
| Aciit | 139 | 109(78%) | 30(22%) | 1.3 | 1.3 |
| Adungosi | 67 | 58(87%) | 9(13%) | 0.8 | 0.0 |

### Sporozoite Rates

A total of 644 samples were tested for sporozoite infectivity rate. The overall mean positivity rate was 1.71%. This varied among species with sporozoite rates of 5.6% (n=5/89), 1.2% (n=6/505) and 2.0% (n=1/50) within *An*. *funestus s.s.*, *An*. *gambiae s.s* and *An. arabiensis*, respectively.

### Human Biting Rates

The human biting rate (HBR) was determined across the study area, with an estimated 2.95 mosquitoes per person per night. Substantial variation in HBR was noted among the villages, with the highest rates being observed in Okaereret (5.9 mosquitoes/person/night), Koruruma (5.2), and Kwangamor (4.2). In contrast, lower rates were documented in Aciit (1.3) and Adungosi (0.8). These variations were found to be statistically significant (P < 0.001) (Table 4).

### Entomological Inoculation Rates (EIR)

The entomological inoculation rate (EIR) was calculated for the study period, with an overall rate of 5.8 infectious bites per person being determined. When analyzed by species, *An. gambiae s.s.* contributed 1.1 infectious bites per person, while *An. funestus* s.s. accounted for 0.81 infectious bites per person for four months. Analysis at complex level showed that *Anopheles gambiae s.l* had the highest EIR compared to *An. funestus s.l*, 4.2 versus 1.7 infective bites/person during the study period respectively. EIR varied across villages, with the highest rate being recorded in Koruruma (19.1 infectious bites/person), followed by Kwangamor (6.7). Lower rates were observed in Okaereret (1.5) and Aciit (1.3), while no infectious bites were recorded in Adungosi (Table 4).

### Relationship Between Malaria Test Positivity Rates and Age, Anophelines Mean densities, HBR and EIR

Using a multilevel logistic modeling approach of village- and individual-level factors as predictors of malaria test positivity. The null model showed an intraclass-correlation coefficient (ICC) of 0.14, suggesting that 14% of the variance in positive malaria test is accounted for by factors at the village level (between-village differences). Several significant associations were identified between the entomological indices and malaria test positivity. A positive association was established between vector density and malaria positivity, with an odds ratio of 1.12 (95% CI: 1.08-1.16, P < 0.0001). Similarly, a positive association was determined between human biting rate and malaria positivity (OR: 1.37, 95% CI: 1.19-1.57, P < 0.0001). A slight negative association was observed with age (OR: 0.999, 95% CI: 0.998-0.9997, P = 0.013), suggesting that younger individuals (under 5 years) were marginally more likely to test positive for malaria. No significant associations were observed between EIR and malaria positivity (OR: 1.04, 95% CI: 0.96-1.13, P = 0.319) or between gender and malaria positivity (OR: 1.01, 95% CI: 0.85-1.21, P = 0.888) (Table 5). For the cross-level interaction, the effect of entomological factors on malaria test positivity did not differ significantly with individual age (OR 1.000, 95% CI .999 - 1.000, *P* = 0.731). Individuals had a 12%, and 37% increase in being infected with malaria when mean density of mosquito and human biting rate increases by one unit respectively.

**Table 5:** Relationship between malaria test positivity rates and *Anopheles* mean densities, HBR, Age, EIR and Gender.

| Variable | OR (95% CI) | P-Value |
| --- | --- | --- |
| Anopheline Mean Density | 1.12, (1.08 - 1.16) | 0.0001 |
| Human Biting Rate (HBR) | 1.37, (1.19 - 1.57) | 0.0001 |
| Age | 0.999, (0.998 - 0.9997) | 0.013 |
| EIR | 1.04, (0.96 - 1.13) | 0.319 |
| Gender | 1.01, (0.85 - 1.21) | 0.888 |

## DISCUSSION

This study demonstrates strong associations between specific entomological indices and malaria test positivity rates in western Kenya, providing critical evidence for optimizing surveillance strategies in high-transmission settings. The significant associations between vector density (OR 1.12, 95% CI: 1.08-1.16) and human biting rate (OR 1.37, 95% CI: 1.19-1.57) with malaria test positivity align with findings from similar high-transmission areas. Recent studies in Ethiopia [12], Tanzania [21], São Tomé and Príncipe [22] have similarly documented strong correlations between vector abundance and disease transmission, attributable to increased human-vector contact probability in areas with higher mosquito densities [13]. The particularly strong association with HBR reflects the dominance of highly anthropophilic *An. gambiae* s.s. and *An. funestus* in the study area [23], consistent with findings from other East African endemic regions where HBR provides direct measurement of human exposure to vector bites [24] [25].

The lack of significant association between EIR and malaria positivity (OR: 1.04, 95% CI: 0.96-1.13) contrasts with some previous studies [26] but can be explained by several factors. Ongoing vector control interventions, particularly widespread ITN use, may have reduced sporozoite rates as demonstrated in western Kenya intervention studies [27]. Additionally, the four-month study duration may have limited ability to capture the full EIR-transmission relationship [28], while complex immunity development in endemic areas can influence the relationship between infectious bites and clinical malaria [29].

The significant spatial heterogeneity in both entomological indices and malaria test positivity across villages aligns with recent comprehensive analyses of malaria transmission patterns in western Kenya [30], [31], where substantial micro-geographic variation has been documented within similar ecological zones [32][33]. The observed heterogeneity can be attributed to local environmental characteristics, including altitude and land use patterns [33], variations in vector control coverage and effectiveness, and socio-economic factors, all of which have been shown to impact local transmission patterns [34].

The finding that younger individuals (under five years) were slightly more likely to test positive for malaria is consistent with established patterns of age-dependent malaria risk in endemic areas. This relationship has been documented [35] explaining that children between 5-14 generally have higher infection rates (prevalence) because they have historically been low priority in malaria control activities (ITNs) and they often have irregular sleeping patterns which means they are often not under a net. The slight negative association with age (OR: 0.999, 95% CI: 0.998-0.9997) follows established age-dependent malaria risk patterns in endemic areas [36], where children under five years show higher clinical incidence due to limited acquired immunity.

The strong predictive value of vector density and HBR measurements, obtainable through relatively simple sampling methods, can be leveraged to optimize surveillance in resource-limited settings. This approach is supported by recent work demonstrating successful implementation of simplified entomological monitoring for routine surveillance [37]. However, while community-based entomological surveillance presents opportunities for cost-effective expansion of data collection [11][38], recent evaluations in western Kenya highlight challenges with accuracy of mosquito identification by community collectors, requiring continued investment in training and quality assurance [11]. Given the WHO’s emphasis on strengthened surveillance systems and the recent global increase in malaria cases (263 million in 2023 compared to 252 million in 2022) [1], integrating entomological indices into routine malaria monitoring can enhance early warning systems and guide targeted interventions. Recent experiences from Côte d’Ivoire demonstrate how entomological monitoring data can effectively drive decision-making for appropriate vector control interventions, including stratified ITN distribution strategies [39]. The observed spatial heterogeneity supports targeted interventions, an approach shown to be cost-effective [40].

The four-month study duration may not have captured seasonal transmission variations, and broader geographical coverage would strengthen regional applicability. Reliance on health facility records may underestimate true disease burden, a common limitation in surveillance studies [41]. Future studies should consider expanding to diverse eco-epidemiological zones as demonstrated in recent regional assessments [31] [30], which enable more generalizable conclusions for intervention planning.

## CONCLUSION

Vector density and human biting rate serve as reliable, easily measurable predictors of malaria transmission intensity that can optimize surveillance strategies in high-endemicity settings. The observed spatial heterogeneity supports targeted interventions, while the strong associations between readily obtainable entomological indices and malaria positivity suggest their utility as cost-effective proxy indicators for transmission monitoring. These findings support the integration of simplified entomological surveillance into routine malaria programs, potentially reducing surveillance costs while maintaining effectiveness in tracking transmission patterns and guiding control efforts.

## Data Availability

All relevant data are within the manuscript and its Supporting Information files.

## Acknowledgement

Special thanks to the Busia County Health Department for their kindness during facility data extraction. We appreciate the residents of the study area for their acceptance during the collection of entomological data.

## Author contribution

Conceptualization: BO, MO, BA, EO. Formal analysis: BO, VM, MO. Funding acquisition: EO. Investigation: BO, EO. Methodology: BO, VM, MO. Project administration: EO. Resources: JG, NA, JPG, EO. Supervision: EO, DM, BA, Writing – original draft: BO, EO, VM. Writing – review & editing: EO, JG, BA, NA, VM.

## References

[1] WHO, World malaria World malaria report report. 2023. [Online]. Available: https://www.wipo.int/amc/en/mediation/%0Ahttps://www.who.int/teams/global-malaria-programme/reports/world-malaria-report-2023

[2] M. Indicator and K. Indicators, “Kenya,” 2020.

[3] P. Klepac, S. Funk, T. D. Hollingsworth, C. J. E. Metcalf, and K. Hampson, “Six challenges in the eradication of infectious diseases,” Epidemics, vol. 10, pp. 97–101, 2015, doi: 10.1016/j.epidem.2014.12.001.

[4] B. Bakhiyi et al., “Public health contributions of entomological surveillance of West Nile virus (WNV) and other mosquito-borne arboviruses in a context of climate change.,” Can Commun Dis Rep, vol. 50, no. 9, pp. 294–304, Sep. 2024, doi: 10.14745/ccdr.v50i09a02.

5. “9789241512978-eng”.

[6] PMI, “President’s Malaria Initiative: Malaria Operational Plan for Kenya 2019,” pp. 1–75, 2019, [Online]. Available: https://www.pmi.gov/docs/default-source/default-document-library/malaria-operational-plans/fy19/fy-2019-kenya-malaria-operational-plan.pdf?sfvrsn=3

[7] E. Chanda, “Malaria Vector Surveillance and Insecticide Resistance Monitoring and Management,” Malar Control Elimin, vol. 4, no. 1, 2015, doi: 10.4172/2470-6965.1000e130.

[8] L. Sedda et al., “Improved spatial ecological sampling using open data and standardization: An example from malaria mosquito surveillance,” J R Soc Interface, vol. 16, no. 153, Apr. 2019, doi: 10.1098/rsif.2018.0941.

9. W. Health Organization, “Guideline WHO Guidelines for malaria - 14 March 2023,” 2023. [Online]. Available: http://apps.who.int/bookorders.

[10] T. L. Russell, R. Farlow, M. Min, E. Espino, A. Mnzava, and T. R. Burkot, “Capacity of National Malaria Control Programmes to implement vector surveillance: a global analysis,” Malar J, vol. 19, no. 1, Dec. 2020, doi: 10.1186/s12936-020-03493-1.

[11] B. Abong’o et al., “Evaluation of community-based vector surveillance system for routine entomological monitoring under low malaria vector densities and high bed net coverage in western Kenya,” Malar J, vol. 22, no. 1, p. 203, Jul. 2023, doi: 10.1186/s12936-023-04629-9.

[12] T. Gari et al., “Malaria incidence and entomological findings in an area targeted for a cluster-randomized controlled trial to prevent malaria in Ethiopia: results from a pilot study,” Malar J, vol. 15, no. 1, p. 145, Dec. 2016, doi: 10.1186/s12936-016-1199-4.

[13] B. Abong et al., “Impact of indoor residual spraying with pirimiphos-methyl (Actellic 300CS) on entomological indicators of transmission and malaria case burden in Migori County, western Kenya,” pp. 1–14, 2020, doi: 10.1038/s41598-020-61350-2.

[14] E. O. Ochomo et al., “Evaluation of the protective efficacy of a spatial repellent to reduce malaria incidence in children in western Kenya compared to placebo: study protocol for a cluster-randomized double-blinded control trial (the AEGIS program),” pp. 1–21, 2022.

[15] W. Health Organization, Training module on malaria control: Malaria entomology and vector control. Guide for participants. 2013. [Online]. Available: https://www.who.int/malaria

[16] M. Coetzee, “Key to the females of Afrotropical Anopheles mosquitoes (Diptera: Culicidae),” Malar J, pp. 1–20, 2020, doi: 10.1186/s12936-020-3144-9.

[17] J. A. Scott, W. G. Brogdon, and F. H. Collins, “Identification of single specimens of the Anopheles gambiae complex by the polymerase chain reaction,” Am J Trop Med Hyg, vol. 49, no. 4, pp. 520–529, 1993, doi: 10.4269/ajtmh.1993.49.520.

[18] “Wirtz RA 1987 (Sporozoite ELISA).pdf”.

[19] MOH, “REPUBLIC OF KENYA MINSTRY OF HEALTH KENYA MALARIA VECTOR SURVEILLANCE OPERATIONAL GUIDELINES (2020-2024) DIVISION OF NATIONAL MALARIA PROGRAMME,” 2020.

[20] “Malaria entomology and vector control Learner’s Guide World Health Organization HIV/AIDS, Tuberculosis and Malaria Roll Back Malaria,” 2003.

[21] G. F. Killeen, N. J. Govella, Y. P. Mlacha, and P. P. Chaki, “Suppression of malaria vector densities and human infection prevalence associated with scale-up of mosquito-proofed housing in Dar es Salaam, Tanzania: re-analysis of an observational series of parasitological and entomological surveys,” Lancet Planet Health, vol. 3, no. 3, pp. e132–e143, Mar. 2019, doi: 10.1016/S2542-5196(19)30035-X.

[22] J. Pinto et al., “Malaria in São Tomé and Principe: parasite prevalences and vector densities,” Acta Trop, vol. 76, no. 2, pp. 185–193, Sep. 2000, doi: 10.1016/S0001-706X(00)00100-5.

[23] J. I. Odero et al., “Early morning anopheline mosquito biting, a potential driver of malaria transmission in Busia County, western Kenya,” Malar J, vol. 23, no. 1, p. 66, Mar. 2024, doi: 10.1186/s12936-024-04893-3.

[24] T. Degefa, A. K. Githeko, M.-C. Lee, G. Yan, and D. Yewhalaw, “Patterns of human exposure to early evening and outdoor biting mosquitoes and residual malaria transmission in Ethiopia,” Acta Trop, vol. 216, p. 105837, Apr. 2021, doi: 10.1016/j.actatropica.2021.105837.

[25] T. S. Churcher et al., “Probability of Transmission of Malaria from Mosquito to Human Is Regulated by Mosquito Parasite Density in Naïve and Vaccinated Hosts,” PLoS Pathog, vol. 13, no. 1, p. e1006108, Jan. 2017, doi: 10.1371/journal.ppat.1006108.

[26] D. L. Smith, J. Dushoff, R. W. Snow, and S. I. Hay, “The entomological inoculation rate and Plasmodium falciparum infection in African children,” Nature, vol. 438, no. 7067, pp. 492–495, Nov. 2005, doi: 10.1038/nature04024.

[27] J. E. Gimnig et al., “The Effect of Indoor Residual Spraying on the Prevalence of Malaria Parasite Infection, Clinical Malaria and Anemia in an Area of Perennial Transmission and Moderate Coverage of Insecticide Treated Nets in Western Kenya,” PLoS One, vol. 11, no. 1, p. e0145282, Jan. 2016, doi: 10.1371/journal.pone.0145282.

[28] L. S. Tusting, T. Bousema, D. L. Smith, and C. Drakeley, “Measuring Changes in Plasmodium falciparum Transmission,” 2014, pp. 151–208. doi: 10.1016/B978-0-12-800099-1.00003-X.

[29] I. Rodriguez-Barraquer et al., “Quantification of anti-parasite and anti-disease immunity to malaria as a function of age and exposure,” Elife, vol. 7, Jul. 2018, doi: 10.7554/eLife.35832.

[30] D. Mategula and J. Gichuki, “Understanding the fine-scale heterogeneity and spatial drivers of malaria transmission in Kenya using model-based geostatistical methods,” PLOS Global Public Health, vol. 3, no. 12, pp. 1–14, 2023, doi: 10.1371/journal.pgph.0002260.

[31] G. Zhou et al., “Malaria transmission heterogeneity in different eco-epidemiological areas of western Kenya: a region-wide observational and risk classification study for adaptive intervention planning,” Malar J, vol. 23, no. 1, pp. 1–11, 2024, doi: 10.1186/s12936-024-04903-4.

[32] A. Kamau, P. Mogeni, E. A. Okiro, R. W. Snow, and P. Bejon, “A systematic review of changing malaria disease burden in sub-Saharan Africa since 2000: comparing model predictions and empirical observations,” BMC Med, vol. 18, no. 1, p. 94, Dec. 2020, doi: 10.1186/s12916-020-01559-0.

[33] C. O. Oduma et al., “Altitude, not potential larval habitat availability, explains pronounced variation in Plasmodium falciparum infection prevalence in the western Kenya highlands,” PLOS Global Public Health, vol. 3, no. 4, p. e0001505, Apr. 2023, doi: 10.1371/journal.pgph.0001505.

[34] S. P. Kigozi et al., “Associations between urbanicity and malaria at local scales in Uganda,” Malar J, vol. 14, no. 1, p. 374, Dec. 2015, doi: 10.1186/s12936-015-0865-2.

[35] J. T. Griffin, N. M. Ferguson, and A. C. Ghani, “Estimates of the changing age-burden of Plasmodium falciparum malaria disease in sub-Saharan Africa,” Nat Commun, vol. 5, no. 1, p. 3136, Feb. 2014, doi: 10.1038/ncomms4136.

[36] R. Ranjha, K. Singh, R. K. Baharia, M. Mohan, A. R. Anvikar, and P. K. Bharti, “Age-specific malaria vulnerability and transmission reservoir among children,” Global Pediatrics, vol. 6, no. July, p. 100085, 2023, doi: 10.1016/j.gpeds.2023.100085.

[37] C. H. Sikaala et al., “A cost-effective, community-based, mosquito-trapping scheme that captures spatial and temporal heterogeneities of malaria transmission in rural Zambia,” Malar J, vol. 13, no. 1, pp. 1–13, 2014, doi: 10.1186/1475-2875-13-225.

[38] US President’s Malaria Initiative, “Niger - The U.S. President’s Malaria Initiative VectorLink Niger Project,” 2023, [Online]. Available: https://stacks.cdc.gov/view/cdc/146401

[39] B. L. Kouassi et al., “Entomological monitoring data driving decision-making for appropriate and sustainable malaria vector control in Côte d’Ivoire,” Malar J, vol. 22, no. 1, pp. 1–15, 2023, doi: 10.1186/s12936-023-04439-z.

[40] I. A. Quakyi et al., “Targeted community based interventions improved malaria management competencies in rural Ghana,” Glob Health Res Policy, vol. 2, no. 1, pp. 1–10, 2017, doi: 10.1186/s41256-017-0048-5.

[41] C. Lourenço et al., “Strengthening surveillance systems for malaria elimination: a global landscaping of system performance, 2015–2017,” Malar J, vol. 18, no. 1, p. 315, Dec. 2019, doi: 10.1186/s12936-019-2960-2.

